# Automated MRI Lung Segmentation and 3D Morphological Features for Quantification of Neonatal Lung Disease

**DOI:** 10.1101/2021.08.06.21261648

**Authors:** Benedikt Mairhörmann, Alejandra Castelblanco, Friederike Häfner, Vanessa Pfahler, Lena Haist, Dominik Waibel, Andreas Flemmer, Harald Ehrhardt, Sophia Stoecklein, Olaf Dietrich, Kai Foerster, Anne Hilgendorff, Benjamin Schubert

**Affiliations:** Institute of Computational Health, Helmholtz Zentrum München, 85764 Neuherberg, Germany, Member of the German Center for Lung Research (DZL); Institute for Lung Health and Immunity and Comprehensive Pneumology Center, Helmholtz Zentrum München, Germany, Member of the German Center for Lung Research (DZL); Center for Comprehensive Developmental Care (CDeC^LMU^) at the interdisciplinary Social Pediatric Center, Dr. von Hauner Children’s Hospital, Hospital of the Ludwig-Maximilian University, Munich, Germany; Department of Neonatology, Perinatal Center, Hospital of the Ludwig-Maximilian University, Munich, Germany; Department of Radiology, Hospital of the Ludwig-Maximilian University, Munich, Germany; Department of Mathematics, Technical University of Munich, 85748 Garching bei München, Germany; Department of General Pediatrics & Neonatology, Justus-Liebig-University, Giessen, Germany, Member of the German Center for Lung Research (DZL); Institute of AI for Health, Helmholtz Zentrum München, 85764 Neuherberg, Germany

**Keywords:** Bronchopulmonary Dysplasia, Chronic Lung Disease, Preterm Infant, Lung Segmentation, Lung Magnetic Resonance Imaging, BPD Severity Prediction, Deep Learning, Lung Imaging Biomarkers, Lung Topology

## Abstract

The diagnosis of neonatal respiratory diseases is currently based on clinical criteria. However, lung structural information is generally lacking due to the unavailability of routinely applicable, radiation-free imaging tools as well as the time-consuming, often non-standardized manual analysis of imaging data. Increased efficiency, comparability and accuracy in image quantification is needed in this patient cohort as pulmonary complications determine immediate and long-term survival.

We therefore developed an ensemble of deep convolutional neural networks to perform lung segmentation in magnetic resonance imaging (MRI) sequences obtained in premature infants near term (n=107), with subsequent reconstruction of the 3-dimensional neonatal lung and estimation of MRI lung descriptors for volume, shape, surface, and signal intensity distribution.

Annotation of lung segments in quiet-breathing MRI for infants with and without Bronchopulmonary Dysplasia (BPD) was achieved by development of a deep learning model reaching a volumetric dice score (VDC) of 0.908 and validated in an independent cohort (VDC 0.880), thereby matching expert-level performance while demonstrating transferability, robustness towards technical (low spatial resolution, movement artifacts) and lung disease grades. MRI lung descriptors presented relevant correlations with lung lesion scores and enabled the separation of neonates with and without BPD (AUC 0.92±0.016), mild vs severe BPD (AUC 0.84±0.027), and single level prediction of BPD severity (AUC 0.75±0.013).

Our work demonstrates the potential of AI-supported MRI markers as a diagnostic tool, characterizing changes in lung structure in neonatal respiratory disease while avoiding radiation exposure.

## INTRODUCTION

The preterm and term neonate postnatally faces the development of acute lung injury with the significant potential to evolve into a chronic disease. With the diagnostic process still solely relying on clinical observation, occasional chest radiography, and late-stage pulmonary function, the application of radiation-free, sensitive imaging strategies and their standardized assessment would critically inform the diagnostic process by adding structural information while allowing for comparability and reproducibility with the goal to implement personalized treatment and monitoring strategies [1–3].

The realization of this currently unmet clinical need is especially challenging in the most vulnerable cohort of preterm infants [1,4,5], burdened with a high incidence of chronic lung disease, *i*.*e*., bronchopulmonary dysplasia (BPD). Here, the low sensitivity and diagnostic value of conventional chest radiography and the limitations of Computed Tomography (CT) due to radiation exposure [6,7] resulted in the exploration of alternative imaging techniques such as Magnetic Resonance Imaging (MRI). Supported by the advantage of the combined assessment of the central nervous system and lung abnormalities, quantitative information and prognostic value of lung MRI for the diseased neonatal lung are currently being explored in infants [8–10], with few studies addressing the MRI assessment of structural changes in the BPD lung [11,12].

In the neonatal lung, MRI is technically challenged by small subject sizes, lower spatial resolution, and sensitivity to infant motion, resulting in blurring, ghosting, and other image artifacts [13]. These conditions demand expert knowledge to obtain qualitative and quantitative measurements from the acquired pulmonary images [9,11] while affecting inter-rater concordances. The subsequently reduced standardization limits high-throughput MRI-based monitoring in neonatal lung disease.

We, therefore, developed a deep learning (DL) based model to enable robust and standardized analysis of lung MRIs in preterm neonates with and without chronic lung disease (BPD) acquired during quiet-breathing near-term age. We combined recent advances in computational methods [4,14,15], *i*.*e*., convolutional neural networks (CNN), to improve the applicability and robustness of deep learning (DL) methods for performing MRI lung segmentation in preterm infants. The obtained lung segmentations were used to compute MRI-based 3-dimensional (3D) lung descriptors including descriptors for shape, surface, and volume. We combined the lung volumetric and structural descriptors to improve disease classification and adequate characterization of BPD, where pre- and postnatal insults provoke a range of structural abnormalities in the immature lung resulting in insufficient gas exchange [1].

## METHODS

### Study Cohort

We prospectively enrolled 107 preterm infants with gestational age (GA) 27±2.1 weeks at birth, from the clinical study Attention to Infants @ Respiratory Risks, with and without later development of BPD at two study sites to perform 3 Tesla Lung MRI near term (GA 37±5.8) during quiet-breathing after informed parental consent (Perinatal Center LMU Munich n=86; EC LMU #195–07; Perinatal Center UKGM Giessen (n=21; ECUKGM #135–12) (**Fig. 1A**).

**Figure 1.**
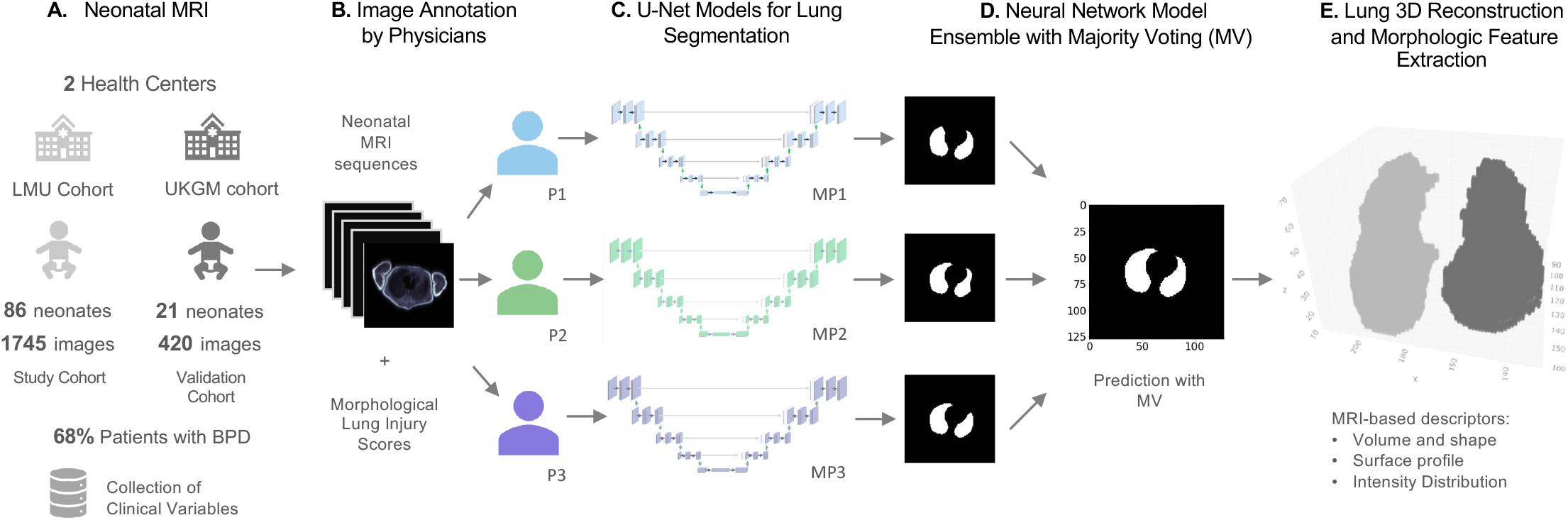
MRI-based Neonatal Lung Volume Analysis Pipeline. **(A)** Clinical study including preterm infants with and without BPD, free-breathing neonatal MRI was taken at Gestational Age (GA) 37 +-5.8 weeks. **(B)** Manual MRI annotation of the lung performed by three trained physicians (P1, P2, P3). MRI morphological injuries (e.g. emphysema, fibrosis, ventilation inhomogeneity) were scored by two trained physicians. **(C-D)** U-Net deep learning models (MP1, MP2, MP3) were trained for lung segmentation, a final lung-mask prediction is calculated with an ensemble of the models through majority voting (MV). **(E)** Lung volume 3D reconstruction and automated calculation of 78 morphologic 3D feature extraction for each lung.

MRI sequences were acquired in unsedated infants (LMU study cohort) and under light sedation with chloral hydrate (30-40 mg/kg (orally), UKGM validation cohort) during quiet sleep in room air. In total, 73 participants were diagnosed with BPD and classified into three severity grades: mild (n=42; requirement of supplemental oxygen for 28 days, no need for oxygen supplementation at 36 weeks PMA), moderate (n=11; requirement of supplemental oxygen for 28 days and oxygen supplementation <FiO_2_ 0.30 at 36 weeks PMA), and severe (n=20; requirement of supplemental oxygen for 28 days and oxygen supplementation >FiO_2_ 0.30 at 36 weeks PMA and/or positive pressure ventilation/continuous positive pressure), based on the NIH consensus definition summarized by Jobe et. al. [1], whereas n=34 infants did not develop BPD. Clinical data acquisition was performed for patients from both cohorts, however, four subjects had to be excluded from the regression analysis due to missing clinical variables (n=103, **Table 1**). Infant lung function testing (ILFT) was performed including tidal breathing analysis and bodyplethsymographic functional residual capacity at 36 weeks GA in n=32 preterm infants [16].

**Table 1.** Patient and Clinical Information of the Preterm Neonatal Cohort (n=103)

| Clinical Variable<br>(Average ±SD) | All<br>($n=103$) | No BPD<br>($n=33$) | BPD Mild<br>($n=39$) | BPD<br>Moderate<br>($n=11$) | BPD<br>Severe<br>($n=20$) |
| --- | --- | --- | --- | --- | --- |
| Gestational Age (weeks) | 26.96<br>$\pm 2.12$ | 29.09<br>$\pm 1.43$ | 26.20<br>$\pm 1.48$ | 25.69<br>$\pm 2.06$ | 25.62<br>$\pm 1.43$ |
| Birth Weight (g) | 908.25<br>$\pm 304.58$ | 1206.21<br>$\pm 292.39$ | 829.74<br>$\pm 182.77$ | 641.82<br>$\pm 177.85$ | 716.25<br>$\pm 154.31$ |
| *Body Size (cm) | 34.38<br>± 4.02 | 38.38<br>±3.23 | 33.26<br>± 2.92 | 31.06<br>±2.43 | 31.76<br>±2.25 |
| APGAR Score - 5 min | 7.71<br>± 1.40 | 8.06<br>±1.00 | 7.87<br>±1.10 | 7.36<br>±2.38 | 7.00<br>±1.59 |
| †Early Onset Infection | No (N=80),<br>Yes (N=23) | No (N=29),<br>Yes (N=4) | No (N=30),<br>Yes (N=9) | No (N=7),<br>Yes (N=4) | No (N=14),<br>Yes (N=6) |
| Administration of postnatal corticosteroids | No (N=61),<br>Yes (N=42) | No (N=28),<br>Yes (N=5) | No (N=22),<br>Yes (N=17) | No (N=6),<br>Yes (N=5) | No (N=15),<br>Yes (N=5) |
| Oxygen Supplementation (days) | 47.30<br>± 43.18 | 5.18<br>±7.72 | 45.56<br>±21.23 | 81.55<br>±30.96 | 101.35<br>±40.78 |
| Mechanical Ventilation (days;<br>invasive and non-invasive) | 48.22<br>±26.93 | 19.91<br>±15.62 | 52.51<br>±13.74 | 66.55<br>±19.20 | 76.50<br>±21.08 |

\*Linear BMI imputation performed for 18 body sizes. †Early Onset Infection as defined by Sherman et al. [17].

### MRI Protocols and Annotations

MRI axial images were obtained (GA 37±5.8) using a T2-weighted half-Fourier-acquired single-shot fast spin-echo (HASTE) sequence with an echo time (TE) of 57 ms, providing sufficient T2-weighted signal and contrast for neonatal lung structural assessment [18]. Spatial resolution was 1.3×1.9 mm^2^ in plane with a slice thickness of 4 mm and 0.4 mm slice gap (**SI 2**).

Manual lung annotation in all MRI sequences was performed independently by three experienced physicians (one senior radiologist, and two late-stage medical students trained in image analysis). The software ITK-SNAP [19] was used to collect the manual segmentations. Pseudonymization of images and clinical information was performed to guarantee blinded analysis. MRI sequences were automatically cropped to 128×128 pixels for model training.

### Morphological MRI Physician Scores

Standardized image analysis was performed by two independent radiologists through scoring of lung morphology addressing characteristic structural changes of BPD [37]. Scoring variables were defined as follows, ‘*interstitial enhancement score’* indicative of the typical lung tissue fibrosis, reflects a distinctive representation of interstitial structures, with thickening of broncho-vascular bundles. The caudo-cranial (CC) and anterior-to-posterior (AP) ‘*AP or CC gradient scores*’ illustrate differences in signal intensities over all lung quadrants showing ventilation inhomogeneity. The ‘*emphysema score*’ quantified the presence of emphysema, with reduced signal intensity, rarefied lung vasculature, hyperexpansion, mosaic pattern of lung attenuation, presence of bullae or blebs. The ‘*atelectasis score*’ indicates partial collapse of a region of the lung, showing consolidation with increased signal intensity. The ‘*airway accentuation score*’ was evaluated based on increased signal intensity in the respiratory ducts and airway wall thickness in relation to airway diameter.

For every score, a semi-quantitative five-point Likert scale was used: a score of ‘1’ represented normal findings, *i*.*e*., the absence of any abnormality, while a score of ‘5’ represented maximum pathology. To achieve a high level of standardization, we virtually segmented the lung into four quadrants. Scoring was performed separately for each variable for the right and left lung and in coronal and axial as well as in sagittal images to allow for the detection of regional differences.

### Deep-Learning MRI Lung Segmentation Model

We trained a set of U-Net CNN models [20] to perform 2D lung segmentation on the collected neonatal MRI scans, with each model based on the manual annotations of a different physician (**Fig. 1B-C**) and combined them through pixel-wise majority voting (MV) to an ensemble model (**Fig. 1D**). A 3D representation of the left and right lung was used to calculate volumetric features that describe the shape, surface, and MRI-intensity distributions (**Fig. 1E**).

U-Net models produce a latent representation of the image by processing it through convolutional layers in a contracting path and through a path of up-convolutional layers, with skipped connections at each level, returning a high-resolution binary pixel-wise segmentation map of the image. Our U-Net architecture has four down and four up-convolutional blocks and a fifth intermediate convolutional block. Batch normalization was included after every building block of the U-Net and a Dropout Layer. Detailed architecture and hyperparameters are available in **SI 3, Table S1 and Table S2**, code available in https://github.com/SchubertLab/NeoLUNet. The Instant-DL framework, which is designed to efficiently train U-Net segmentation models for medical imaging applications, was adapted for our study [21]. Optimization was performed with Adam [22].

To generate unbiased training and performance estimates for the study cohort, a set of k models were trained in a leave-one-patient-out (LOPO) cross-validation scheme, that is, for each k^th^ model, the data of the k^th^ participant is used only for validation. Additionally, MRI sequences from the UKGM cohort were used for validation of the model trained with all the sequences from the LMU study cohort. Lung segmentation performance was measured by aggregating pixels from all the slices in the MRI sequence and calculating the volumetric dice coefficient (VDC), defined as 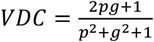with p being the predicted positive-class pixels and *g* being the ground truth For each MRI sequence, the segmentation generated by one of the DL models (*e*.*g*., MP1), previously trained with the manual annotations obtained from one physician (*e*.*g*., P1) was compared with each ground-truth of the remaining manual segmentations (*i*.*e*., P2 and P3). We report the average model performance across these comparisons. The average inter-rater VDC concordance between physicians (*e*.*g*., P1 vs each manual annotation P2 and P3) was calculated as a reference for the model performance. The MV ensemble model prediction was evaluated by comparing its performance against a ground truth generated with all manual annotations (P1, P2, P3) aggregated through pixel-wise majority voting.

### MRI-Lung Volumetric and Morphologic Descriptors

After using the deep convolutional neural networks to perform lung segmentation in MRI sequences, we subsequently reconstructed the 3-dimensional neonatal lung, and estimated MRI lung descriptors for volume, shape, surface, and signal intensity distribution.

The 3D representation of the lung was created by thresholding the predicted lung masks and generating voxels with the DICOM pixel-spacing metadata from the MRI sequences. Voxels that were connected in the 3D space were used to form the final volumetric representations of each lung. Lungs were oriented to a common reference frame for all patients. Left and right lungs were then determined by the x-coordinates of the components’ centroids, enabling the extraction of side-specific lung features for each patient (**SI 4**).

A set of 78 MRI-based lung features were obtained as quantitative descriptors for the morphology of the left and right lung. The 3D descriptors are based on the work described in the work by Waibel et al. [24]. Additionally, we propose MRI-intensity-spacial-distribution features to further reflect lung injury, which were calculated using *Scikit-Image 0*.*19*.*2* [25]. The descriptors proposed can be grouped within the following representative categories: ***Volume and shape features*** (n=38), describing lung volumes and volume ratios, major and minor axis lengths, centroids, inertias and moments of inertia for each 3D axis. ***Surface descriptors*** (n=10), quantifying surface area, surface roughness and surface convexity. ***Intensity distribution features*** (n=30), that include intensity weighted centroids, distances of the MRI intensity-weighted centroids to the non-weighted centroids and descriptive statistics of the central tendency and dispersion of pixel intensities in the lung (**S5** and **Table S3, SI)**.

### BPD Severity Prediction Models

Beyond the explanatory analysis, machine learning (ML) regression models were used to predict the severity of BPD, as well as the primary BPD indicators (days of respiratory support and days of oxygen supplementation), using combinations of three groups of explanatory variables: 78 lung volumetric and morphologic features (***V***), four patient (***P***) features (*i*.*e*., GA, birth weight, body size, gender), and three clinical parameters (***C***) (*i*.*e*., 5 min APGAR score, early-onset infection, steroid treatment).

Random Forest (RF) [26] and Logistic regression models (LR) with Elastic Net [27] regularization were trained to perform binomial classification of two scenarios (no BPD vs BPD; no/mild vs moderate/severe BPD) and multinomial classification (no BPD, mild, moderate, and severe BPD), using *scikit-learn v*.*1*.*1*.*1 [28]*.

A nested cross-validation scheme was implemented to find the best model hyperparameters with a randomized-search (**SI 6, Table S4**). The average performance of the models was estimated with 10 random repetitions of the nested cross-validation scheme. A stratified 5-fold train-test split was used for the inner and outer cross-validation loops.

Our training pipeline also allowed the inclusion of univariate feature selection and dimensionality reduction methods. Principal Component Analysis (PCA), when applied, involved the lung descriptors only, thus keeping the interpretability of the remaining features. Ultimately, a set of LR and RF models were trained both with and without the feature selection methods, with the aim of finding the best performing combination of model, hyperparameters, and features (see **SI 6**).

For prediction of the continuous BPD indicator, regression models (*i*.*e*., Poisson and RF), were trained to predict the days of required respiratory support and days of oxygen supplementation, using the same nested cross-validation and feature selection schemes.

## RESULTS

### Deep Learning Enables Robust MRI Neonatal Lung Segmentation and Lung Volume Calculation Across Study Sites and Disease Grades

The key of establishing MRI-based diagnostics in clinical practice is a robust and standardized analysis protocol. To this end, we developed a deep convolutional neural network for MRI-Lung segmentation and reconstruction in very premature infants near term.

The DL lung segmentation models achieved high VDC performance (MP1=0.890±0.041, MP2=0.878±0.042, MP3=0.872±0.043) equivalent to the inter-rater segmentation concordances (P1=0.875±0.032, P2=0.881±0.034, and P3=0.879±0.035) (**Fig. 2B**), with average VDC differences of less than 0.016 points, demonstrating the capacity of the model for abstracting the MRI lung representation from the training set. Moreover, differences in the average VDC performance between sites were below 0.0286 points for all models, indicating transferability across independent cohorts and model generalizability.

**Figure 2.**
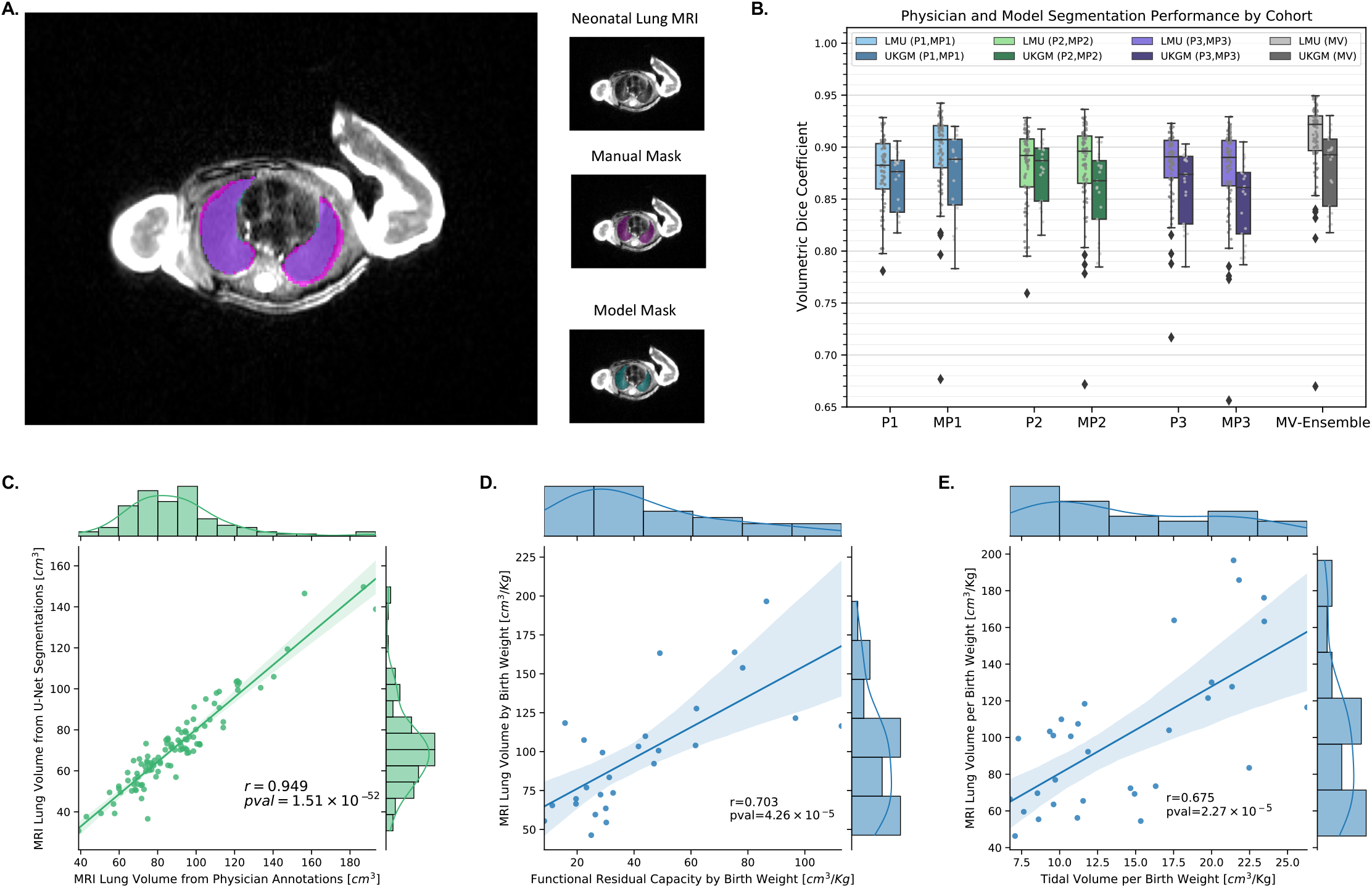
Lung Segmentation and Lung Volume Analysis. **(A)** MRI Lung segmentation sample with manual annotation and ML model generated lung masks. **(B)** Lung segmentation performances for manual physician-based lung annotations (P1, P2, P3), U-Net models (MP1, MP2, MP3) and ensemble model with majority voting (MV), results separated for the LMU study cohort and UKGM validation cohort. **(C)** Estimated MRI lung volume from the U-Net model segmentations vs estimated lung volume from manual segmentations (n=107). **(D)** Functional residual capacity per birth weight vs MRI model-based lung volume per birth weight (n=27). **(E)** Tidal volume per birth weight vs MRI model-based lung volume per birth weight (n=32).

The MV ensemble model showed the highest VDC when compared to the rater’s consensus (**Fig. 2B**), with an average VDC of 0.902±0.039 (study cohort=0.908±0.039 and validation cohort=0.880±0.036), confirming human-level accuracy of the AI-based segmentation method for quiet-breathing neonatal lung MRI. VDC scores per MRI sequence are available (**Table S5, SI)**. Image quality was analyzed as a confounding factor for segmentation performance (**Fig. S1A, SI**), independent scores per sequence (1=high quality, 2=medium quality, 3=low quality) were on average 1.7 for the study cohort and 2.3 for the validation cohort. A significant effect was found for both manual segmentations (Kruskal-Wallis P1, P2, P3, p-values=[1.24×10^−6^, 1.14×10^−5^, 4.21×10^−8^], n=107) and the models accordingly (Kruskal-Wallis MP1, MP2, MP3, MV, p-values=[1.53×10^−7^, 7.14×10^−7^, 4.67×10^−8^, 2.53×10^−7^], n=107), lower MRI quality resulted in lower segmentation performances.

Model segmentation robustness was tested for different disease conditions, *i*.*e*., the presence of BPD-characteristic structural changes, and showed no significant differences for segmentation performance between BPD severity grades (**Fig. S1B, SI**) (Kruskal-Wallis MP1, MP2, MP3, MV, p-values=[0.30, 0.20, 0.55, 0.48], n=107). Similarly, model segmentation performance was not significantly affected by different types of lung injury, including: interstitial enhancement, emphysema, atelectasis, ventilation inhomogeneities, and airway accentuation (**Fig. S2A-F, SI**), showing comparable performances to the manual annotations in the same pathologic conditions.

The correlation of the resulting MRI lung volumes per patient, computed using DL-based vs manual lung segmentations, was evaluated as another indicator of segmentation accuracy (**Fig. 2C**). A significantly high correlation (Pearson, r=0.949, p-value=1.51×10^−52^, n=107) was found, indicating that the precision of the DL ensemble model enabled a robust downstream estimation of the lung volumes, even for sequences with low image quality (Pearson, r=0.950, p-value=1.65×10^−09^, n=18).

The DL-based lung volumes per patient were also validated against MRI-independent volume estimators from ILFT, a significant positive correlation was observed between the DL lung volumes normalized by birthweight vs the functional residual capacity normalized by birth weight (**Fig. 2D**, r=0.703, p-value=4.26×10^−5^, n=27), as well as the tidal volume normalized by birth weight (**Fig. 2E**, r=0.675, p-value=2.27×10^−5^, n=32).

### MRI Lung Features Correlate with BPD Severity Indicators and Morphological Lesion Scores

Our explanatory analysis demonstrated relevant correlations of the MRI lung features against indicators of BPD severity and lung morphological injury scores (**Fig. 3**; **Table S6, SI)**.

**Figure 3.**
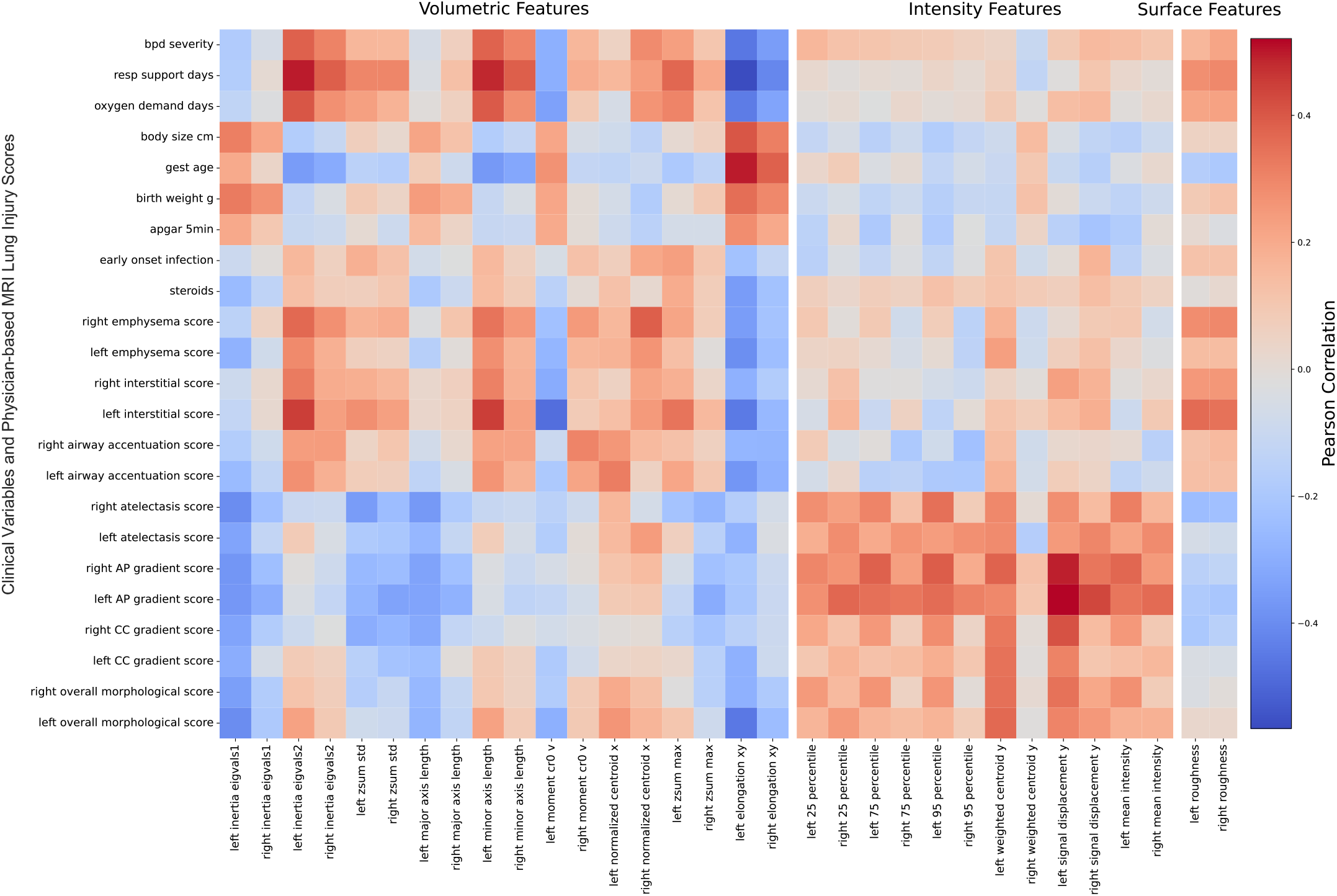
Correlation Matrix of MRI-based Morphological Features vs Clinical Variables and Lung Injury Scores. Exploratory analysis of the MRI-based 3D morphological features that presented highest pearson correlation with clinical variables or MRI physician-based lung injury scores. MRI 3D features are grouped by feature type (Volumetric, Intensity and Surface). AP=Antero-Posterior, CC=Caudo-Cranial.

In concordance with the pathophysiology of the disease, clinical features related to GA presented good correlations with BPD severity levels (Pearson; gestational-age: r=-0.518, p-value=1.63×10^−^, birth-weight: r=-0.571, p-value=6.97×10^−8^; body-size: r=-0.588, p-value=2.37×10^−8^). We found that MRI lung features such as the MRI-based lung volume normalized by birth weight, also showed a good correlation with BPD (Pearson; r=0.562, p-value=6.52×10^−10^), and allowed an accurate discrimination between BPD severity grades (Kruskal-Wallis, k=42.17, p-value=3.68×10^−9^, n=103), with significant differences between four comparisons of BPD severity levels (Wilcoxon–Mann–Whitney U test with Bonferroni correction: no BPD vs mild BPD, no BPD vs moderate BPD, no BPD vs severe BPD, and mild BPD vs severe BPD, k=[236, 25, 60, 217], p-values=[2.54×10^−5^, 1.41×10^−4^, 4.56×10^−6^, 3.44×10^−2^], n=103; **Fig. 4A**). Moreover, high correlations were observed between MRI lung volume normalized by birth weight with days of mechanical ventilation (r=0.738, p-value=5.54×10^−19^, n=103; **Fig. 4B**), and days of oxygen supplementation (r=0.622, p-value=2.39×10^−12^, n=103; **Fig. 4C**). These findings are in line with previous observations reporting elevated lung volumes in severe BPD cases [9,11].

**Figure 4.**
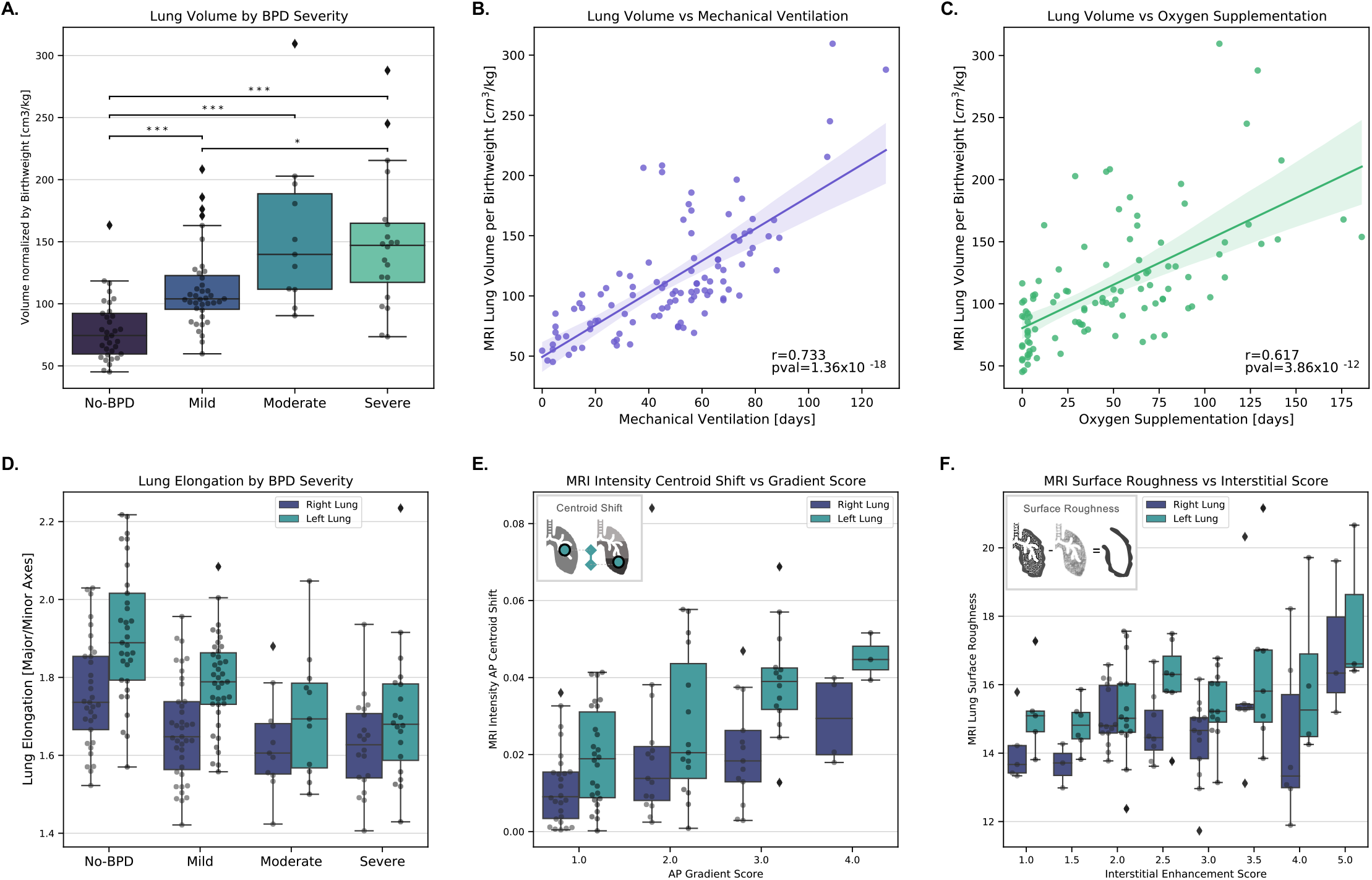
MRI Lung Volumetric Features against BPD Severity and Lung Injury Scores. **(A)** Distribution of the predicted lung volume normalized by birth weight against BPD severity grades. (n=103, *p-values for Wilcoxon–Mann–Whitney U-test with Bonferroni correction). **(B**) Lung volume normalized by birth weight vs days of mechanical ventilation (n=103). **(C)** Lung volume normalized by birthweight vs days of oxygen supplementation (n=103). **(D)** Lung Volumetric Elongation (Major Axis / Minor Axis) by BPD Severity (n=103). **(E)** MRI lung intensity Antero-Posterior (AP) centroid shift vs AP gradient Score indicating ventilation inhomogeneity (n=58). **(F)** MRI lung volumetric surface roughness vs interstitial lung injury score indicating fibrosis (n=58).

Lung elongation (*i*.*e*., major over minor lung axis length) also demonstrated relevance for describing BPD severity levels (Pearson, left-lung: r=-0.46, p-value=1.02×10^−6^ ; right-lung, r=-0.35., p-value=2.80×10^−4^; n=103) (**Fig. 4D)** and days of mechanical ventilation (Pearson, left-and-right-lungs: r=0.465, p-value=1.98×10^−12^, n=103), details in (**Table S7, SI 7)**.

Next, we explored the correlation of lung 3D morphologic features with physician-based morphological lung injury scores. In particular, the lung surface roughness, which measures lung surface irregularities by subtracting the raw 3D lung shape from its gaussian-smoothed version, showed a significant positive correlation with the interstitial lung injury score that indicates interstitial remodeling (Pearson; left-lung: r=0.362, p-value=5.20×10^−3^, n=58) (**Fig. 4F)**. Also, the MRI AP-centroid shift, which describes the distance in [cm] between the intensity-weighted centroid and the centroid with uniform pixel intensity distributions, showed a positive correlation with the AP intensity-gradient score (Pearson; left-lung: r=0.521, p-value=2.75×10^−5^; right-lung: r=0.337, p-value=9.67×10^−3^; n=58) (**Fig. 4E**), corroborating the potential to be used as a quantitative descriptor for ventilation inhomogeneities.

### MRI Lung Features Demonstrate Predictive Performance for BPD Severity Classification

To evaluate the clinical potential of the MRI-based lung volumetric analysis in the preterm neonate, we tested the performance of ML models for BPD severity prediction with GA, and Patient (P), Clinical (C), and MRI-lung (L) features.

For the binary BPD classification, a high accuracy for separating BPD and no BPD cases was found when using only GA (AUC 92.06%), PC features (AUC 92.14%), or PCL features (AUC 91.67%) (**Table 2)**, which reflects the driving force of immaturity for the development of lung injury and BPD. In the binary separation of no and mild from moderate and severe BPD cases, we found that the inclusion of MRI-lung features as explanatory variables (PCL) improved the average AUC predictive performance by 8.36 % points when compared to only GA and 2.29 % points when compared with the predictions with PC variables (**Fig. 5A**).

**Table 2.** BPD Severity Prediction by Feature Groups.

|  | AUC [%] | PCL | PC | GA | L |
| --- | --- | --- | --- | --- | --- |
| **Binary: No BPD vs BPD (all severity levels)** | Average<br>± SD | 91.67<br>±1.57 | 92.14<br>±1.14 | 92.06<br>±0.92 | 74.71<br>±1.23 |
|  | *Best model* | *LR (UFS)* | *LR* | *LR* | *LR (PCA-l)* |
| **Binary: No/Mild BPD vs Moderate/Severe BPD** | Average<br>± SD | **84.11**<br>**±2.66** | 81.82<br>±2.21 | 75.75<br>±1.34 | 75.20<br>±2.94 |
|  | *Best model* | *LR (PCA-l, UFS)* | *RF* | *LR* | *LR* |
| **Multinomial: No BPD, BPD Mild, Moderate and Severe.** | Average<br>± SD | 75.21<br>±1.34 | 75.71<br>±1.23 | 74.84<br>±2.04 | 60.75<br>±2.27 |
|  | *Best model* | *LR (UFS)* | *RF* | *LR* | *LR* |
| **Multinomial: No BPD, BPD Mild, Moderate and Severe.**<br>**GA Bin [25.4-28.6]** | Average<br>± SD | **67.60**<br>**±2.60** | 62.62<br>±3.72 | 54.65<br>±3.79 | 60.23<br>±5.17 |
|  | *Best model* | *LR (UFS)* | *RF* | *LR* | *RF(PCA-l)* |

**PCL**=Patient, clinical and lung descriptors, **PC**=Patient and clinical descriptors, **GA**=Gestational age, **L**=78 MRI lung volumetric and morphological descriptors, **LR**=Logistic Regression, **RF**=Random Forest, **PCA-l**=Principal component analysis for lung features. **UFS**=Univariate feature selection. **P**=Patient descriptors (gestational age, birth weight, body size, gender), **C**=Clinical Parameters (APGAR 5 min score, early-onset infection, postnatal steroids treatment). Average AUC scores and standard deviations (SD) across 10 repetitions of the nested cross validation.

**Figure 5.**
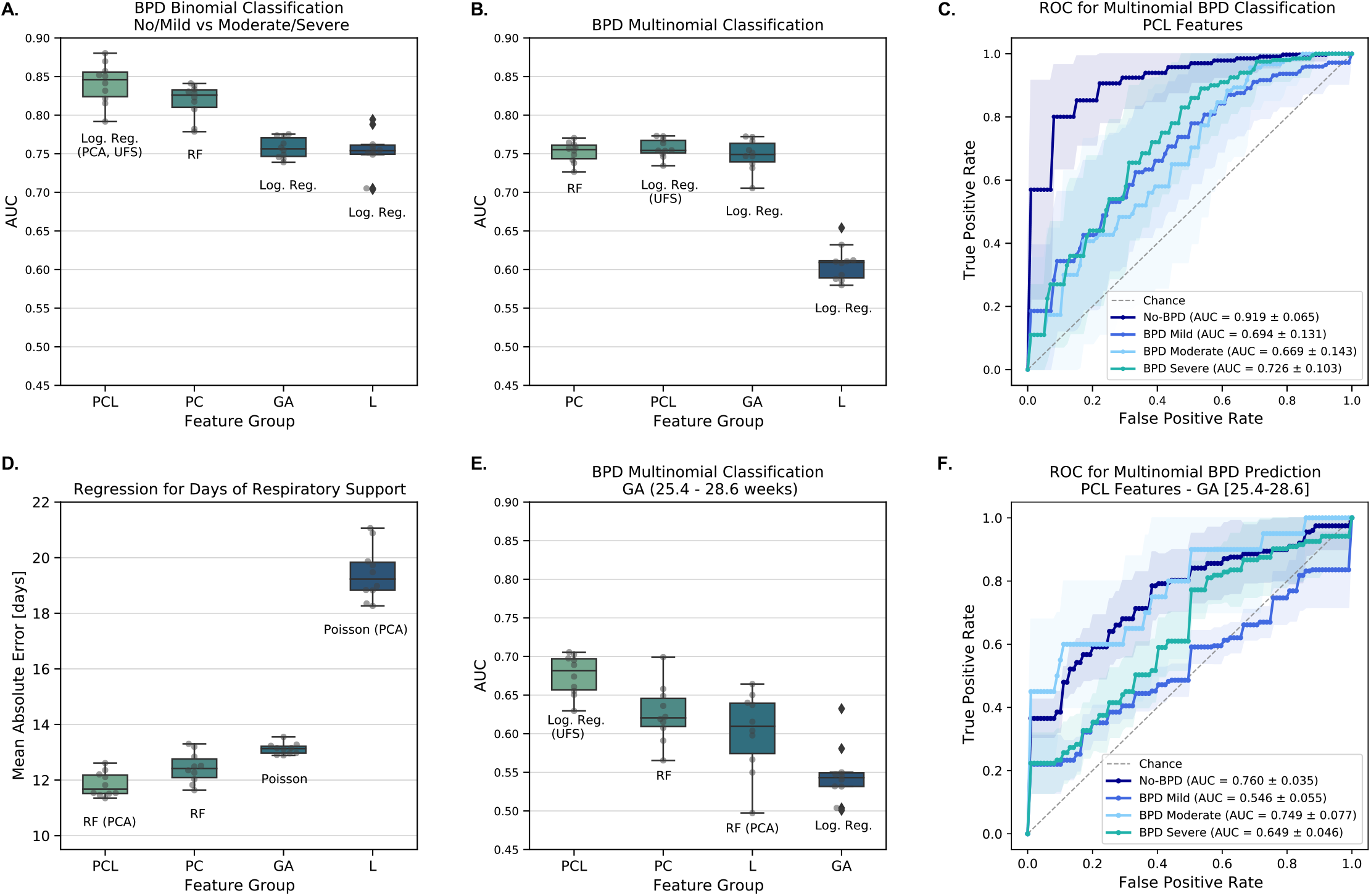
BPD Prediction - Best Performing Models by Feature Group. **(A)** BPD binomial classification performance (No/Mild vs. Moderate/Severe). **(B)** BPD multinomial classification performance (No, Mild, Moderate, Severe). **(C)** BPD multinomial ROC for best model with PCL features. **(D)** Regression performance for days of respiratory support. **(E)** BPD Multinomial classification performance for GA (25.4-28.6 weeks) **(F)** BPD multinomial ROC for best model with PCL features with GA (25.4-28.6 weeks). Feature Groups (PCL=Patient, clinical and lung features, PC=Patient and clinical variables, GA=Gestational age, L=78 MRI automated lung volumetric and morphological descriptors). Log. Reg.=Logistic Regression, RF=Random Forest, PCA=Principal component analysis only for lung features. UFS=Univariate feature selection.

The multiclass prediction of BPD severity showed comparable performance for the PCL and PC models, with macro-weighted AUCs of 75.21% and 75.71% respectively, (**Table 2, Fig. 5B**). The individual intra-class performance of the PCL model showed a high score for classification of no BPD vs all BPD levels (AUC=91.9%), followed by severe BPD detection (AUC=72.6%) (**Fig. 5C**).

Furthermore, we investigated the models’ performance in a subgroup of patients (n=50) with a GA between 25.4 to 28.6 weeks established by including the most immature patient without BPD and the most mature patient classified with severe BPD. Within this group of extremely immature infants, GA alone does not sufficiently discriminate disease severity (AUC=54.65%). In contrast, the inclusion of lung morphological features largely improved the separation of BPD grades for the PCL model with an overall performance improvement of 4.98% points above the best PC model, and 12.95% points above the GA model (**Table 2, Fig. 5E**). The average intra-class AUC scores of the PCL model for this particular GA window were [75.97%, 54.65%, 74.90%, 64.91%] for no, mild, moderate, and severe BPD, respectively (**Fig. 5F**).

To understand the contribution of the lung MRI-based features for prediction of BPD severity outcome, we performed a permutation importance analysis on a LR multinomial classification model (*LR + UFS*) trained with all the features (PCL) and data-points (**Fig. S4A, SI**), we found that together with GA and birth weight, 16 MRI-based lung features had importances with interquartile ranges above random chance. Also, we investigated the feature importances of the RF models trained for BPD outcome prediction in every outer fold of the cross-validation (**Fig. S4B, SI**), we observed that after GA, birth weight, and body size, 17 additional MRI-based lung features were part of the top 20 features used for BPD prediction. In both analyses the MRI-based *lung volumes, lung elongation, intensity-centroids, inertias and moments*, showed relevant contribution for BPD classification.

For the continuous indicators of BPD, by the use of RF regression, and Poisson regression, the prediction of days of mechanical respiratory support and oxygen supplementation was evaluated (**Table 3**). For the prediction of days with respiratory support, the regression model that included patient, clinical and MRI-lung features (PCL), achieved the lowest mean absolute error (MAE) with 11.85 days, compared to 12.44 days with the PC features, and 13.12 days with GA-only (**Fig. 5E)**. In the case of prediction of days with oxygen supplementation, the PCL model had similar performance to the PC and GA models, with a MAE of 23.94 days.

**Table 3.** Prediction of Primary BPD indicators by Feature Groups.

|  | MAE [days] | PCL | PC | GA | L |
| --- | --- | --- | --- | --- | --- |
| **Regression: Days of Respiratory Support** | Average<br>± SD | **11.85**<br>**±0.44** | 12.44<br>±0.55 | 13.12<br>±0.20 | 19.43<br>±0.97 |
|  | *Best model* | *RF(PCA-l)* | *RF* | *Poisson* | *Poisson (PCA-l)* |
| **Regression: Days of Oxygen Supplementation** | Average<br>± SD | 23.94<br>±1.00 | 23.42<br>±1.18 | 22.28<br>±0.41 | 33.23<br>±4.03 |
|  | *Best model* | *RF(PCA-l)* | *Poisson* | *Poisson* | *Poisson (PCA-l)* |

MAE=Mean Absolute Error, **PCL**=Patient, clinical and lung descriptors, **PC**=Patient and clinical descriptors, **GA**=gestational age, **L**=78 MRI lung volumetric and morphological descriptors, **RF**=Random Forest, **PCA-l**=Principal Component Analysis for Lung Features. **P**=Patient descriptors (gestational age, birth weight, body size, gender), **C**=Clinical Parameters (APGAR 5 min score, early-onset infection, postnatal steroids treatment). Average AUC scores and standard deviations (SD) across 10 repetitions of the nested cross validation.

## DISCUSSION

We showcased the significant potential of DL models for accurate segmentation of neonatal lung MRI as well as the use of MRI-based lung injury descriptors for the standardized and reproducible assessment of the lung MR images in a high-risk patient cohort.

With low variability and high comparability, the performance of the developed CNN models outweighed manual annotations, thereby demonstrating their significant potential to perform in smallest lung volumes, challenged by motion artifacts and blurring.

Previous studies faced limitations in scalability and sensitivity of MRI lung segmentation in smaller neonatal cohorts. In line with this, Heimann et al. used lung shape-appearance models to perform free-breathing MRI lung segmentation in a cohort of 32 children and reported an average overlap of volumetric ground truth of 85% [29]. Kohlmann et al. achieved a ground truth segmentation overlap of 94% using 3D lung region-growing-based methods in MRIs from 14 adult patients while applying breath-holding maneuvers [30], whereas other adult MRI lung segmentation methods reported VDCs in the range of 82%-86% [31,32]. In contrast, we demonstrated that our ensemble DL model achieved an average performance VDC of 90.2% in the most challenging condition, *i*.*e*., free-breathing neonatal lung MRI obtained in a multi-center setting, thereby comparable or superior to the performance of segmentation models designed for highly controlled acquisition protocols in adult patients. Furthermore, our ensemble model demonstrated robust performance in disease-associated structural changes, indicating the potential of the model to be applicable in various, clinically relevant conditions.

As a result, our automated pipeline enabled the accurate downstream estimation of neonatal lung volumes that significantly correlated with the corresponding volumes abstracted from manual annotations (Pearson, r=0.949). The performance was comparable to previous MRI lung volume extractions in adults (Pearson, r=0.98) [30]. Moreover, our MRI volume estimates correlated with ILFT as a non-image-based validation of the findings obtained.

To showcase the potential for diagnostic use, we demonstrated significant correlations of multiple MRI-lung descriptors with physician-based lung injury scores. For instance, the intensity-weighted centroid shift allowed us to link the deviation of the centroid to the degree of ventilation inhomogeneities. In addition, the correlation between lung surface ‘roughness’ and interstitial enhancement suggested that extensive matrix remodeling in BPD, with concurrent emphysematous and fibroproliferative changes, is reflected in larger 3D surface irregularities, in line with findings in adult lung fibrosis [33].

Previous MRI lung injury features in neonatal cohorts focused on the quantification of lung water content via proton density measurements [34] or examined lung MRI relaxation times [18], thereby lacking information of the lung topology and MRI-intensity distributions. MRI 3D morphological descriptors have the potential to further contribute as interpretable, quantitative markers of lung structural injuries in neonates.

The application of our MRI-supported prediction models for disease stratification demonstrated significant predictive power for BPD classification (AUC 91.67±1.6% no BPD vs BPD), thus exceeding previous radiation-free imaging-supported BPD prediction models (AUC binary prediction performance: 83-86% (lung ultrasound) [8–10], 80% (lung MRI) [18]). The inclusion of MRI-lung features also improved the predictive performance for the separation of more severe BPD cases (AUC 84.11%), thereby stratifying high-risk infants. Moreover, the multinomial classification of BPD with PCL features showed improved performance (AUC 75.21%) when compared to 13 of the 14 clinical BPD outcome prediction models (AUC 54% - 73%), and resembled the model of Ryan et al. (AUC 76%) [35,36], based on the validation by Onland et al. [35]. Our analysis furthermore revealed that MRI morphologic lung features significantly improved BPD classification in more immature infants, in which GA does not sufficiently predict BPD severity.

With the potential of quantifiable MRI lung structural information to improve the precision for identification of BPD cases, our results motivate further research in the extraction of MRI-lung volumetric and morphological descriptors to better guide medical care and interventions as well as long-term monitoring.

For future work, the collection of larger annotated datasets will strengthen the generalizability and performance of the ensemble model while including different conditions of lung pathology. Moreover, the definition of additional lung descriptors and their analysis in longitudinal approaches will be crucial to inform medical decision-making through early prediction of long-term outcome.

Our work contributes to the generation of scientific evidence required to integrate AI-driven, volume-based MRI descriptors that can serve as markers for neonatal lung health in clinical routine taking advantage of a radiation-free imaging technique. The proposed segmentation method and automated extraction of structural measurements from neonatal lung MRI enables the translation of medical expertise to larger-scale applications, including the transferability to health centers that face different expertise levels. Therefore, this approach contributes to the standardized monitoring of critical features in pediatric respiratory disease and improves the comparability and reproducibility in image analysis for standardized follow-up care into adulthood.

## Data Availability

Source code is available at https://github.com/SchubertLab/NeoLUNet

## Data and Source Code Availability

Source code of models for lung segmentation, 3D volume-feature estimations, and regression models can be found at https://github.com/SchubertLab/NeoLUNet Resulting weights of the U-Net models used for segmentation and Models for BPD prediction will be made available at (https://zenodo.org).

## Ethics statement

### Patient consent for publication

Not required.

### Ethics approval

This study was approved by the Ethics Board of the Ludwig-Maximilians University Hospital, Munich, Germany (EC LMU #195-07) and the Ethics Board of the Justus-Liebig-University, Giessen, Germany (EC UKGM #135-12).

## Acknowledgments

We sincerely thank the patients and their families of the AIRR study cohort for their significant contribution to the study by providing the samples.

