## Supplementary material for "Automated MRI Lung Segmentation and 3D Morphological Features for Quantification of Neonatal Lung Disease": SI

† The authors wish it to be known that, in their opinion, the last two authors should be regarded as Joint Last Authors.

**Keywords:** Bronchopulmonary Dysplasia, Chronic Lung Disease, Preterm Infant, Lung Segmentation, Lung Magnetic Resonance Imaging, BPD Severity Prediction, Deep Learning, Lung Imaging Biomarkers, Lung Topology.

**SUPPLEMENTARY INFORMATION**

1. **Methods: Cohort Characteristics**

Preterm infants with a gestational age ≤32 weeks were prospectively included in the study on informed parental consent (Study Site 1, n=86, Perinatal Centre LMU Munich, EC #195–07; Study Site 2, n=21, Perinatal Centre UKGM Giessen, EC #135-12). BPD (mild, moderate and severe) was diagnosed at 36 weeks gestational age. Inclusion of participants for the study started in December 2012 and ended November 2020. Pulmonary function tests were performed according to the guidelines of the American Thoracic and European Respiratory Society.

2. **Methods: Imaging and Annotation Protocols**

Axial images were obtained (GA 37±5.8) with a T2-weighted half-Fourier-acquired single-shot turbo spin echo (HASTE) protocol for lung structural assessment. An ECG-triggered 2D multi-slice single-shot fast spin-echo sequence with an echo time (TE) of 57 ms was used; the repetition time was set to 2 RR intervals.

The T2-weighted sequence with an echo time of 57 ms shows low signal intensities in normal (non-pathological) lung parenchyma, predominantly as an effect of low proton density in the lung. However, in contrast to gradient-echo (GRE) acquisitions with non-ultrashort TEs, there is still a reasonable level of (spinecho-refocused) signal remaining in the lungs. By choosing relatively narrow windowing settings, these images show that the lung signal is significantly higher than the air/background signal outside the subject. T2-weighted lung acquisitions can also generally be well suited for the depiction of lung pathologies such as infiltrates or nodules (Hatabu et al. *https://doi.org/10.1148/radiol.2020201138*).

The spatial MRI resolution was 1.3×1.9 mm² in plane with a slice thickness of 4 mm and 0.4 mm slice gap. Parallel imaging with an acceleration factor of 2 was applied and 2 averages were acquired for each slice.

A total of 107 Pulmonary MRI sequences with 2,165 axial images, with a resolution of 256×192 pixels, were acquired during spontaneous quiet breathing without oxygen supplementation and without invasive or non-invasive respiratory support in supine position swaddled in a vacuum mattress after feeding using neonatal noise attenuators (Minimuffs®, natus® newborn care, Seattle, USA) for hearing protection. Study infants did not exhibit clinical or laboratory signs of infection. Two types of 3T MRI scanners were used (Siemens Skyra for the Perinatal Centre LMU Munich and Siemens Verio for Perinatal Centre UKGM Giessen).

To remove unnecessary background, we first identify the centroid of all pixels that are above the 5% intensity quantile threshold across all slices and then crop all slices to a square of 128×128 pixels centered at the centroid.

Standardized image scoring of lung structural injury was performed by two independent radiologists (i.e., a third-year radiological resident and a radiology fellow).

### 3. Methods: Deep-Learning MRI Lung Segmentation Model

Hyperparameters of the U-Net segmentation models were optimized using grid search for three randomly selected leave-one-patient-out models in a 4-fold cross-validation scheme, the best performances were achieved with 300 training epochs, 0.001 learning rate, using a binary cross-entropy loss for the NN optimization, and applying image augmentations with 0.1 random zoom, 0.1 translations and up to 22.5° random rotations.

Architecture parameters of the UNets are summarized in **Table S1**, and hyperparameters for random search in **Table S2**.

### 4. Methods: MRI-Lung Volumetric Representation

The lung 3D volume was built from the lung segmented 2D masks by generating voxels in the 3D space, each voxel has dimensions of (dx,dy) constituting the distance to the neighboring pixels, and (dz) representing slice thickness and space between the MRI slices, as obtained from the DICOM metadata.

A pipeline to orient all the patients' 3D lung volumetric representations to a common reference frame was developed and applied. The pipeline consists of calculating a convex hull silhouette for each binary 2D lung mask (including left and right lung masks) from the MRI slides; we then calculate the rotation of the minor axis $\theta_i$ for each convex hull.

Afterwards, the overall rotation angle $\theta_{avg}$ of the 3D lung object is found by performing a weighted average of the individual slice rotations $\theta_i$ , weighted by the lung area of the slice $a_i$:

$$\theta_{avg} = \frac{\sum_{slice=i}^{N} \theta_i \cdot a_i}{\sum_{slice=i}^{N} a_i}$$

Rigid rotation of the lung 3D object is applied such that $\theta_{avg}$ is aligned with the x-axis of the new reference frame. The pipeline includes regionprops functions from the *Scikit-Image 0.19.2* library (See code repository).

Ultimately, the left and right separation of the lungs is performed by finding the two largest 3D objects with connected voxels in the 3D space, the left or right designation of the lungs is determined by the x coordinates of the 3D object centroids, enabling the extraction of side-specific lung features for each patient.

### 5. Methods: 3D Lung Morphological Features

MRI-Morphological features were investigated as descriptors of lung disease, based on the topological features proposed by Waibel et al. [24], and measures to quantify the distribution of pixel intensities from the lung 3D representations. The features evaluated fall in three category groups:

- ***Volume and shape features*** (n=38): Total volume, left and right lung volume, left over right volume ratio. Minor axis length, major axis length, lung elongation, normalized centroids (x,y,z), maximum lung height and standard pixel-columns height deviation in the z-dimension, for both left and right lungs. Three eigenvalues of the inertia of each lung's volume. Six descriptors of the moments of inertia per lung.
- ***Surface descriptors*** (n=10): Surface area, surface roughness, gauss surface roughness, surface convexity, and 3D boundaries per lung.
- ***Intensity distribution features*** (n=30): MRI signal intensity weighted centroids (x,y,z), the distance of the MRI intensity-weighted centroids to the non-weighted centroid (x,y,z), maximum, minimum, median, standard deviation, and percentiles of the lung pixel intensities (5th, 25th, 50th, 75th, 95th).

A description of the complete set of features can be found in **Table S3**.

### 6. Methods: BPD Severity Prediction Models

Logistic regression (LR) with elastic-net and random forest (RF) models for BPD severity prediction (Binomial, Multinomial and Regression) were trained and evaluated in a nested cross-validation (CV) scheme, with 5-fold splits in both the inner and outer k-folds. The best model hyperparameters were found using a randomized search algorithm in the inner fold and model performance was validated in the outer fold of the CV; ranges and configurations given for the hyperparameter search are described in **Table S4**.

In addition, model performance with and without applying of feature selection methods, namely Principal Component Analysis (PCA) and/or Univariate Feature Selection (UFS), was also evaluated. PCA was used to reduce the dimensionality of only the lung morphological features, and the resulting PCA-components were filtered to include only those which constituted the top (90% or 99%) of the cumulative variance ratio. UFS was also optionally applied for dimensionality reduction of all the input features before model training; the top 20 features of the UFS were selected with the mutual information coefficient (MI).

The different combinations of explanatory feature groups (**PCL**=Patient, clinical and lung MRI-descriptors, **PC**=Patient and clinical descriptors, **GA**=Gestational age, **L**=78 MRI lung MRI-descriptors) and model pipeline configurations (LR or RF, with/without PCA and UFS) were trained and tested each on an individual CV scheme. Only the best performing model configuration for each feature group (best average throughout the outer CV folds) was reported in the main manuscript.

### 7. Results: MRI Lung Features Correlate with BPD Severity

Automated MRI-lung features extracted from each MRI-sequence are available in **Table S6.** Our exploratory analysis showed that MRI-based lung morphological features have significant correlation with BPD severity levels. In **Table S7**, we show the statistical tests (Pearson, Kruskal-Wallis and Wilcoxon–Mann–Whitney U-test with Bonferroni correction) for correlation and paired wise discrimination of BPD severity levels with the two most highly correlated MRI-based morphological variables and clinical variables.

### Supplementary Tables

#### *Table S1:* U-Net Architecture Parameters

| | **Block Description** |
|---|---|
| Convolutional Block 1<br><br>(CNN-1) | CNN Filters = 64, Kernel = 3×3<br>Activation function = LeakyReLU (alpha=0.2)<br>Batch Normalization<br><br>CNN Filters = 64, Kernel = 3×3<br>Activation function = LeakyReLU (alpha=0.2)<br>Batch Normalization |
| Dropout and Max Pooling | Dropout fraction = 0.1<br>Max. Pooling Kernel Size = 2×2 |
| Convolutional Block 2<br>(CNN-2) | CNN Filters = 128, Kernel = 3×3<br>Activation function = LeakyReLU (alpha=0.2)<br>Batch Normalization<br><br>CNN Filters = 128, Kernel = 3×3<br>Activation function = LeakyReLU (alpha=0.2)<br>Batch Normalization |
| Dropout and Max Pooling | Dropout fraction = 0.1<br>Max. Pooling Kernel Size = 2×2 |
| Convolutional Block 3<br>(CNN-3) | CNN Filters = 256, Kernel = 3×3<br>Activation function = LeakyReLU (alpha=0.2)<br>Batch Normalization<br><br>CNN Filters = 256, Kernel = 3×3<br>Activation function = LeakyReLU (alpha=0.2)<br>Batch Normalization |
| Dropout and Max Pooling | Dropout fraction = 0.1<br>Max. Pooling Kernel Size = 2×2 |
| Convolutional Block 4<br>(CNN-4) | CNN Filters = 512, Kernel = 3×3<br>Activation function = LeakyReLU (alpha=0.2)<br>Batch Normalization<br><br>CNN Filters = 512, Kernel = 3×3<br>Activation function = LeakyReLU (alpha=0.2)<br>Batch Normalization |
| Dropout and Max Pooling | Dropout fraction = 0.1<br>Max. Pooling Kernel Size = 2×2 |
| Convolutional Block 5<br>(CNN-5) | CNN Filters = 1024, Kernel = 3×3<br>Activation function = LeakyReLU (alpha=0.2)<br>Batch Normalization<br><br>CNN Filters = 1024, Kernel = 3×3<br>Activation function = LeakyReLU (alpha=0.2)<br>Batch Normalization |
| Dropout and Max Pooling | Dropout fraction = 0.1<br>Max. Pooling Kernel Size = 2×2 |
| Convolutional Block 6 | Up-6 Features: feature size= 512, up-sampling-kernel = 2×2 |

| | |
|---|---|
| (CNN-6) | Concatenation: CNN-4 Features + Up-6 Features<br><br>CNN Filters = 512, Kernel = 3×3<br>Activation function = LeakyReLU (alpha=0.1)<br>Batch Normalization<br><br>CNN Filters = 512, Kernel = 3×3<br>Activation function = LeakyReLU (alpha=0.1)<br>Batch Normalization |
| Convolutional Block 7<br>(CNN-7) | Up-7 Features: feature size= 256, up-sampling-kernel = 2×2<br>Concatenation: CNN-3 Features + Up-7 Features<br><br>CNN Filters = 256, Kernel = 3×3<br>Activation function = LeakyReLU (alpha=0.1)<br>Batch Normalization<br><br>CNN Filters = 256, Kernel = 3×3<br>Activation function = LeakyReLU (alpha=0.1)<br>Batch Normalization |
| Convolutional Block 8<br>(CNN-8) | Up-8 Features: feature size= 128, up-sampling-kernel = 2×2<br>Concatenation: CNN-2 Features + Up-8 Features<br><br>CNN Filters = 128, Kernel = 3×3<br>Activation function = LeakyReLU (alpha=0.1)<br>Batch Normalization<br><br>CNN Filters = 128, Kernel = 3×3<br>Activation function = LeakyReLU (alpha=0.1)<br>Batch Normalization |
| Convolutional Block 9<br>(CNN-9) | Up-9 Features: feature size= 64, up-sampling-kernel = 2×2<br>Concatenation: CNN-1 Features + Up-9 Features<br><br>CNN Filters = 64, Kernel = 3×3<br>Activation function = LeakyReLU (alpha=0.1)<br>Batch Normalization<br><br>CNN Filters = 64, Kernel = 3×3<br>Activation function = LeakyReLU (alpha=0.1)<br>Batch Normalization |
| Output Layer | Activation function = Sigmoid |

#### *Table S2:* U-Net Grid Search Parameters.

| | **Hyperparameters Evaluated with Grid Search** |
|---|---|
| **U-Net for Lung Segmentation** | Epochs = [100, 200, 300, 400]<br>Loss Functions = Mean squared error, Binary cross-entropy, Dice-loss.<br>Learning Rate = [0.001, 0.0001]<br><br>Augmentation = Augmentations were applied with the following configuration (0.1 random zoom, 0.1 translations and up to 22.5° random rotations), training without augmentations was also compared. |

*Table S3:* **3D MRI-based Feature Descriptions**

| | **Feature Name** | **Description** |
|---|---|---|
| **Volume and Shape Features** | total volume [voxels] * | sum of the voxel volumes for the left and right lung |
| | left right volume ratio * | volume of left lung divided by volume of the right lung |
| | number of lung voxels | number of voxels of each lung |
| | major axis length | length of a lung's major axis calculated on a 3D segmentation volume using scikit-image regionprops library. |
| | minor axis length | length of a lung's minor axis calculated on a 3D segmentation volume using scikit-image regionprops library. |
| | normalized centroid z | the z-location of the lung's centroid relative to its height in the z-axis (values in [0,1]) |
| | normalized centroid y | the y-location of the lung's centroid relative to its length in the y-axis (values in [0,1]) |
| | normalized centroid x | the x-location of the lung's centroid relative to its width in the x-axis (values in [0,1]) |
| | zsum std | standard deviation of the heights of all voxel columns in the z-axis |
| | zsum max | maximal height of all voxel columns in the z-axis |
| | moment cr0 v | raw image moments up to the third order calculated using the moments function of scikit image |
| | moment cc0 v | raw image moments up to the third order calculated using the moments function of scikit image |
| | moment cr1 v | raw image moments up to the third order calculated using the moments function of scikit image |
| | moment cc1 v | raw image moments up to the third order calculated using the moments function of scikit image |
| | moment cr2 v | raw image moments up to the third order calculated using the moments function of scikit image |
| | moment cc2 v | raw image moments up to the third order calculated using the moments function of scikit image |
| | inertia eigvals0 | the eigenvalues of the inertia tensor of the image calculated using the inertia_tensor_eigvals function of scikit-image |
| | inertia eigvals1 | the eigenvalues of the inertia tensor of the image calculated using the inertia_tensor_eigvals function of scikit-image |
| | inertia eigvals2 | the eigenvalues of the inertia tensor of the image calculated using the inertia_tensor_eigvals function of scikit-image |
| | elongation | the lung's major axis length divided by its minor axis length |
| **Intensity-Based Features** | mean intensity | mean value of a lung's mri signal values which are scaled to the range [0,1] within the combined lung mask |
| | intensity weighted centroid z | coordinates of the normalized centroid as described above but weighted by scaled mri signal |
| | intensity weighted centroid y | coordinates of the normalized centroid as described above but weighted by scaled mri signal |
| | intensity weighted centroid x | coordinates of the normalized centroid as described above but weighted by scaled mri signal |

| | | |
|---|---|---|
| | intensity std | standard deviation of a lung's mri signal values which are scaled to the range [0,1] within the combined lung mask |
| | max intensity | maximum of a lung's mri signal values which are scaled to the range [0,1] within the combined lung mask |
| | min intensity | minimum of a lung's mri signal values which are scaled to the range [0,1] within the combined lung mask |
| | intensity 5 percentile | percentile values of a lung's mri signal values which are scaled to the range [0,1] within the combined lung mask |
| | intensity 25 percentile | percentile values of a lung's mri signal values which are scaled to the range [0,1] within the combined lung mask |
| | intensity 50 percentile | percentile values of a lung's mri signal values which are scaled to the range [0,1] within the combined lung mask |
| | intensity 75 percentile | percentile values of a lung's mri signal values which are scaled to the range [0,1] within the combined lung mask |
| | intensity 95 percentile | percentile values of a lung's mri signal values which are scaled to the range [0,1] within the combined lung mask |
| | centroid intensity shift z | absolute value of the difference between the z coordinates of the normalized centroid and intensity-weighted centroids |
| | centroid intensity shift y | absolute value of the difference between the y coordinates of the normalized centroid and intensity-weighted centroids |
| | centroid intensity shift x | absolute value of the difference between the x coordinates of the normalized centroid and intensity-weighted centroids |
| **Surface Descriptors** | surface 3D boundary | number of voxels in the intersection of the lung mask and the binary dilation (scikit image function) of the same mask |
| | surface gauss roughness | absolute value of the pointwise difference of the predicted lung array and its gaussian blurred version (scikit image function gaussian_filter used for blurring) |
| | surface area | number of voxels in the predicted lung array's mesh which is interpolated using the marching cubes algorithm (scikit image) |
| | surface roughness | mean squared distance of vertices of the mesh described above and the smoothed mesh which is generated using the humphrey filter |
| | surface convexity | voxel volume of predicted lung array divided by voxel volume of the corresponding convex hull array |

*Features calculated across both lungs (not side-specific)

#### *Table S4:* Randomized Search Parameters for BPD Classification Models.

| | **Hyperparameters Description and Range** |
|---|---|
| **Logistic Regression (Binomial, Multinomial)** | ● The regularization for Logistic Regression models was performed choosing L1, L2 or Elastic-net penalty functions.<br>● Regularization strength parameter *C* was sampled from a uniform logarithmic function with range [0.0001, 10], using scipy.stats.<br>● For elastic-net regularization, the l1/l2 ratio was sampled from a uniform distribution (*loc*=0, *scale* =1) using scipy.stats.<br>● A weighted F1 score was chosen as the metric for performance of the randomized hyperparameter search. |
| **Random Forest** | ● The max. tree depth was sampled from a random uniform distribution of integers [5,100]. |

| | |
|---|---|
| **(Binomial, Multinomial, Regression)** | ● The number of RF estimators was sampled from a random uniform distribution of integers [50, 500]<br>● A weighted F1- score was chosen as the metric for performance of the randomized hyperparameter search in the classification models. |
| **Poisson (Regression)** | ● Regularization strength parameter (*alpha*) was sampled from a uniform distribution (*loc*=0, *scale* =1) using scipy.stats.<br>● The negated value of the mean absolute error was chosen as the metric for performance of the randomized hyperparameter search in the regression models. |

***Table S5:*** **Segmentation Performances for Models and Manual Annotations**

Volumetric Dice Coefficient per MRI sequence for Physicians (P1 vs [P2, P3], P2 vs [P1, P3], P3 vs [P1, P2]) and Models (M1 vs [P2, P3], M2 vs [P1, P3] , M3 vs [P1, P2]) and ensemble model MV vs [P1,P2,P3], can be found in the Supplementary Files.

***Table S6:*** **MRI-Lung Features per Sequence**

Calculated MRI-Lung Features per MRI sequence are available in the supplementary files.

***Table S7 :*** **Exploratory Analysis - Features vs BPD Severity**

| | **Pearson's correlation** | **Kruskal-Wallis** | **Pairwise comparison Wilcoxon–Mann–Whitney (WMW) U-test** (with bonferroni correction) * |
|---|---|---|---|
| **MRI Lung Volume by Birth Weight** [cm3/Kg] | r=0.562<br>p-value=6.52e-10 | k=42.17<br>p-value=3.68e-09 | WMW = [236, 25, 60, 107, 217, 112]<br>p-values=[2.54e-05, 1.41e-04, 4.56e-06, 7.33e-02, 3.44e-02, 1.00]<br>significance=[***, ***, ***, N.S, *,N.S] |
| **MRI Lung Elongation** (*left lung*) | r=-0.460<br>p-value=1.02e-06 | k=26.01<br>p-value=9.48e-06 | WMW = [944, 296, 550, 294, 557, 114]<br>p-values=[6.98e-04, 2.00e-03, 5.63e-05, 6.43e-02, 7.68e-03, 0.885]<br>significance=[**, *, ***, N.S, *, N.S] |
| **Gestational Age** [weeks] | r=-0.587<br>p-value=7.42e-11 | k=48<br>p-value=1.72e-10 | WMW = [1173, 328, 623, 270,470,102]<br>p-values=[2.21e-09, 7.034e-05, 7.42e-08, 0.193, 0.203, 0.772]<br>significance=[***, ***, ***, N.S, N.S, N.S] |
| **Birth Weight** [Kg] | r=0.563<br>p-value=1.17e-07 | k=27.31 p-value=5.07e-06 | WMW=[620, 142, 325, 138, 284, 35]<br>p-values=[9.75e-04, 2.00e-03, 5.00e-05, 0.0437, 0.0639, 0.592]<br>significance=[***, *, ***, N.S, N.S, N.S] |

* Order for the WMW Pairwise BPD Tests (No vs Mild, No vs Moderate, No vs Severe, Mild vs Moderate, Mild vs Severe, Moderate vs Severe). Significance (***=p-value≤ 0.001, **=p-value≤0.01, *=p-value≤0.05, N.S = Not Significant, p-value>0.05).
