## Supplementary material for "Automated MRI Lung Segmentation and 3D Morphological Features for Quantification of Neonatal Lung Disease": Fig. S1

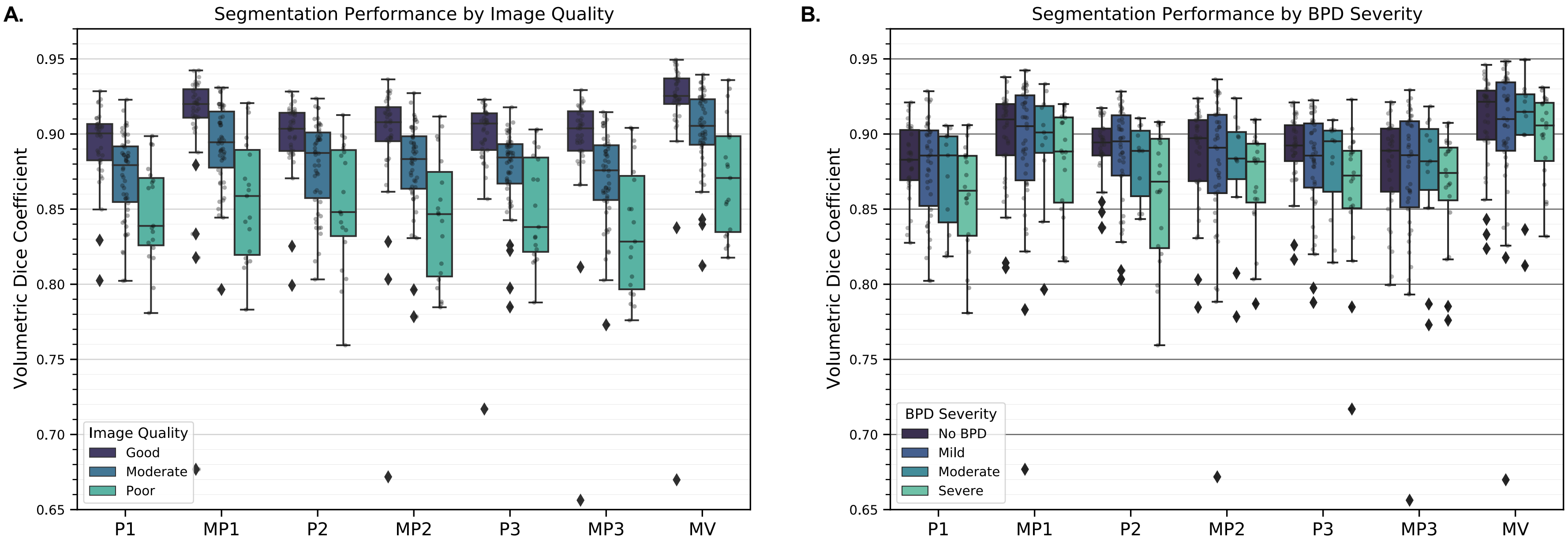


**Figure S1. Lung Segmentation Performance by Image Quality and BPD Severity. (A)** Lung segmentation performances for physician-based lung annotations (P1, P2, P3), U-Net models (MP1, MP2, MP3) and ensemble model with majority voting (MV), results discriminated by image quality. **(B)** Lung segmentation performances discriminated by BPD severity.
