## Supplementary material for "Automated MRI Lung Segmentation and 3D Morphological Features for Quantification of Neonatal Lung Disease": Fig. S2

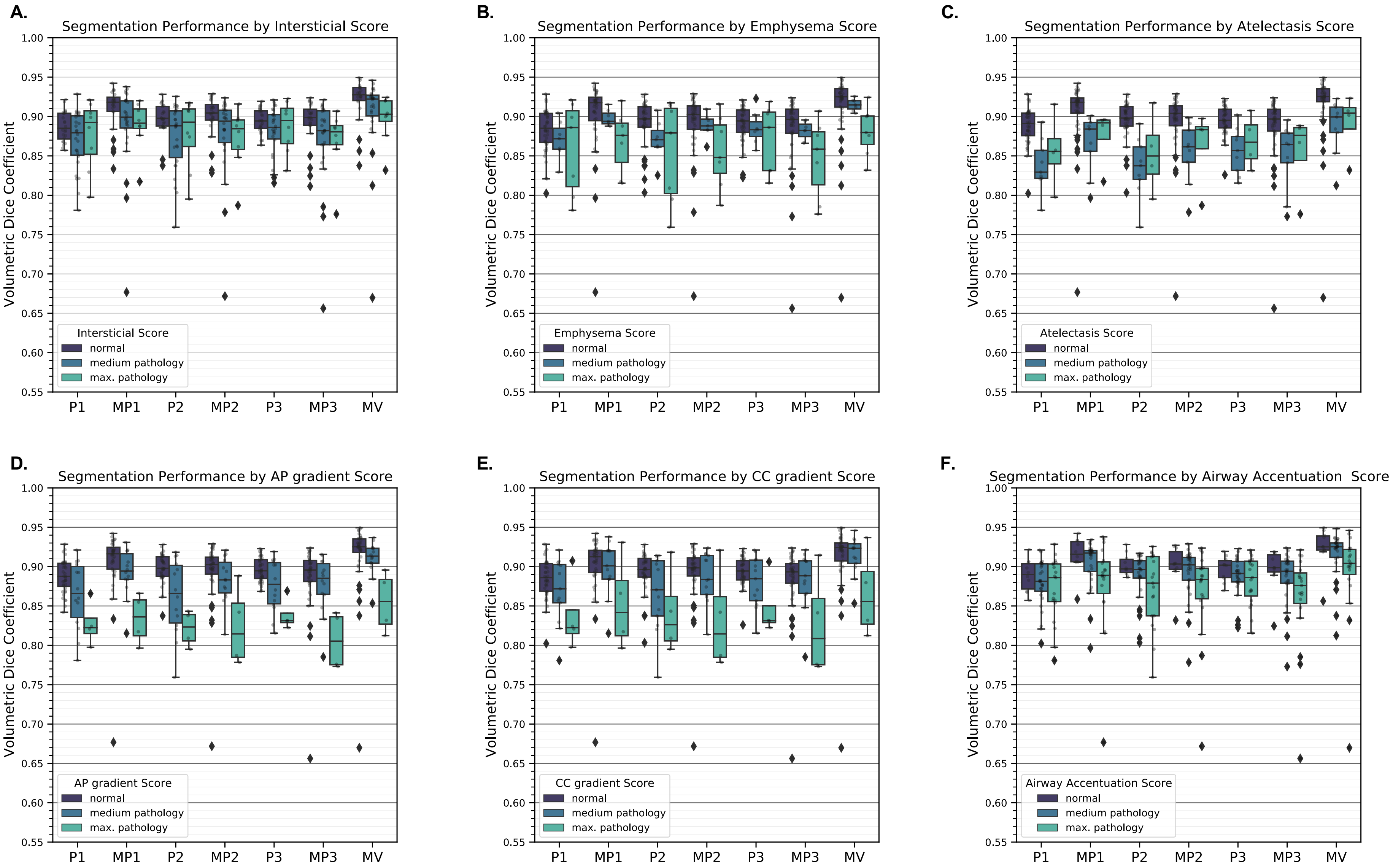


**Figure S2. Lung Segmentation Performance by Lung Lesion Scores.** Lung segmentation performances for physician-based lung annotations (P1, P2, P3), U-Net models (MP1, MP2, MP3) and ensemble model with majority voting (MV), results discriminated by physician-based morphological lesion scores binned by (normal, medium pathology and maximum pathology). **(A)** Segmentation performance vs interstitial Score. **(B)** Segmentation performance vs emphysema Score. **(C)** Segmentation performance vs atelectasis score. **(D)** Segmentation performance vs Antero-Posterior (AP) gradient score. **(E)** Segmentation performance vs Caudal-cranial (CC) gradient score. **(F)** Segmentation performance vs airway accentuation score.
