## Supplementary material for "Automated MRI Lung Segmentation and 3D Morphological Features for Quantification of Neonatal Lung Disease": Fig. S3

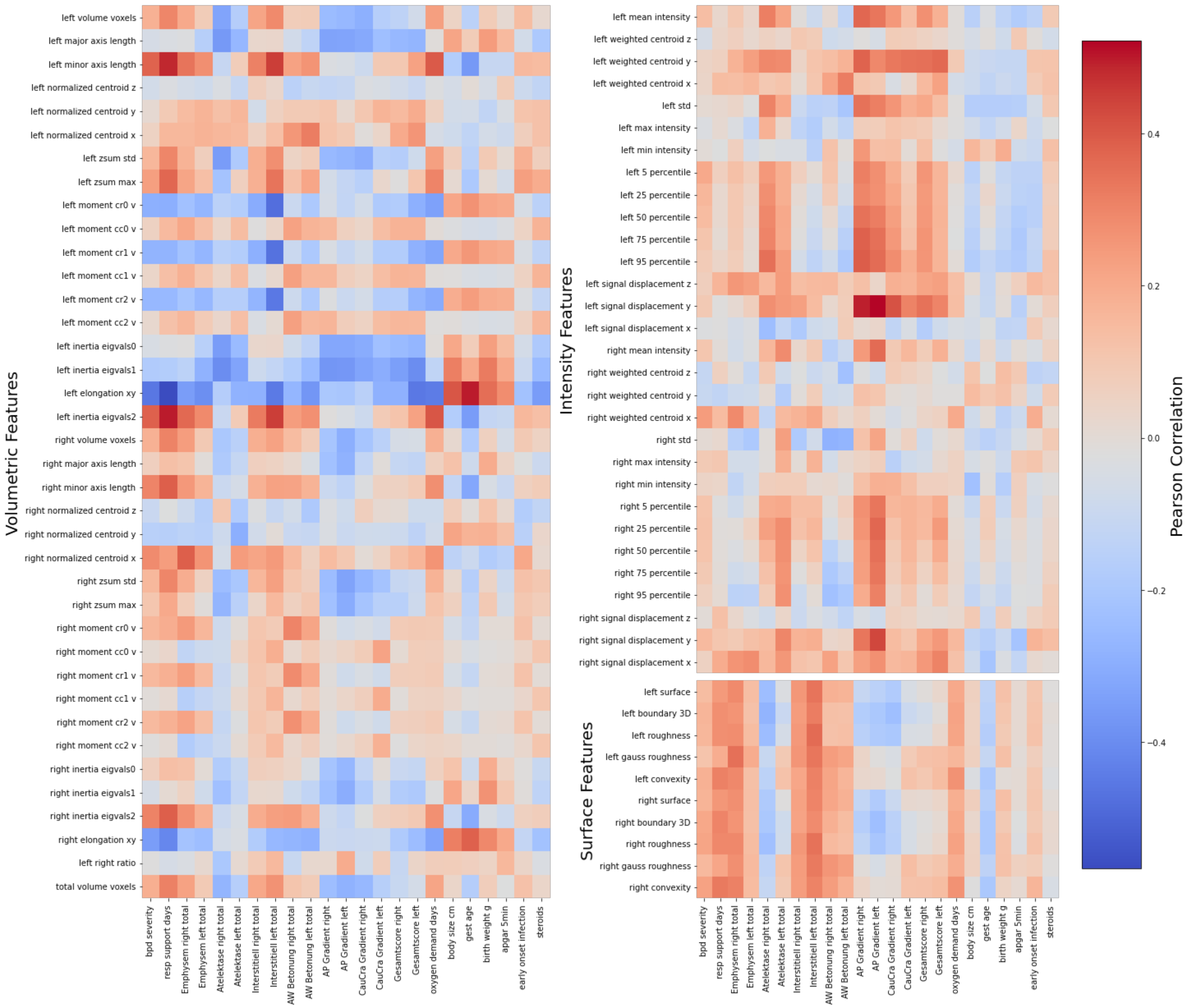


**Figure S3. Correlation Matrix of MRI-based Morphological Features vs Clinical Variables and Lung Injury Scores.** MRI-based 3D morphological features vs clinical variables or MRI physician-based lung injury scores. MRI 3D features are grouped by feature type (Volumetric, Intensity and Surface). AP=Antero-Posterior, CC=Caudo-Cranial.
