## Supplementary material for "Automated MRI Lung Segmentation and 3D Morphological Features for Quantification of Neonatal Lung Disease": Fig. S4

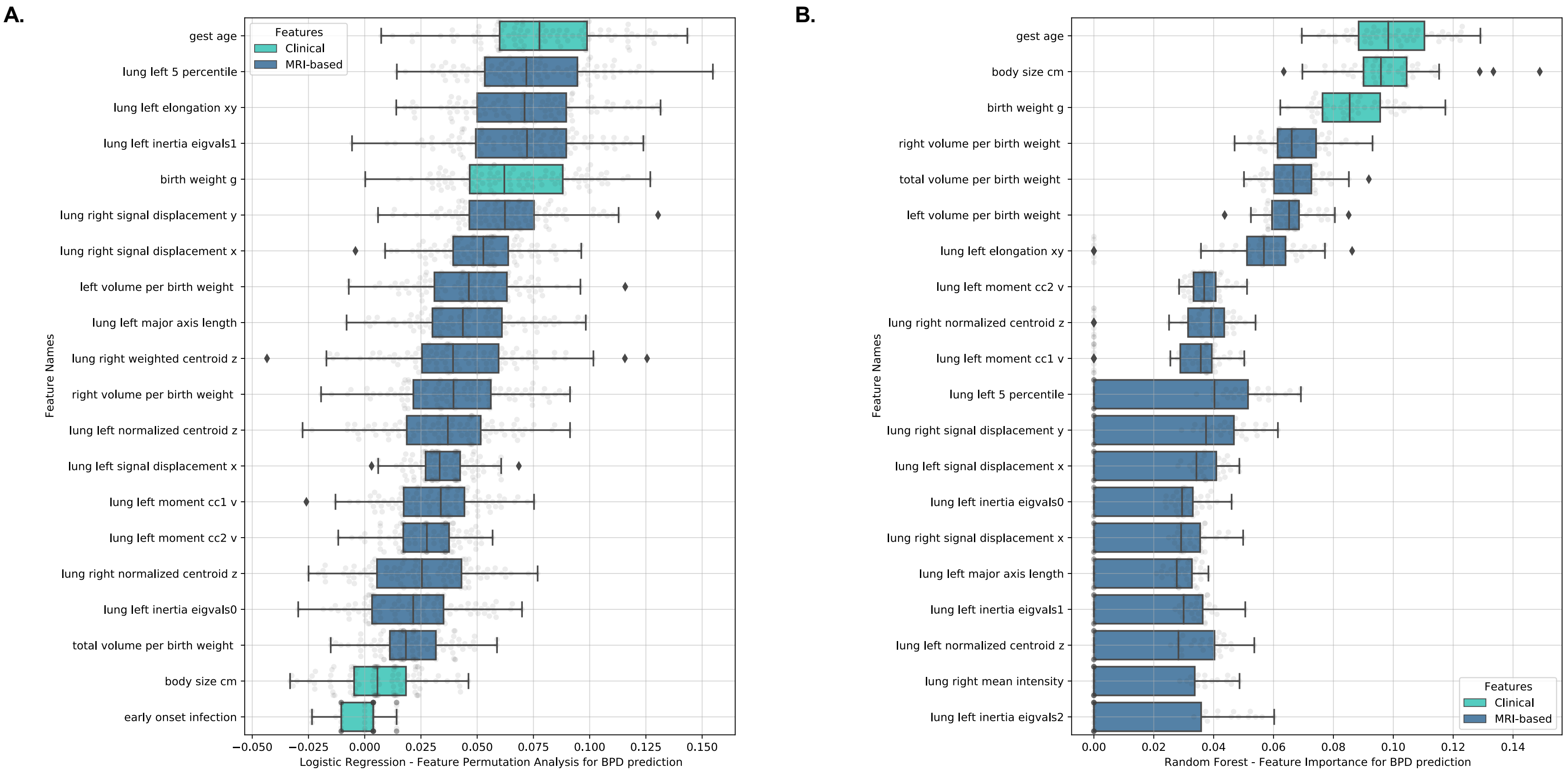


**Figure S4. Feature Importance for BPD Prediction. (A)** Permutation Importance Analysis for the Multinomial Logistic Regression Model (100 repetitions). **(B)** Distribution of feature importance of the Random Forest models for Multinomial Classification in the cross-validation scheme (5 outer folds and 10 repetitions). Explanatory variables for (A) and (B) include Patient, Clinical and MRI-based Lung Features.
